# Sweat lactate sensor for detecting anaerobic threshold in heart failure: a prospective clinical trial (LacS-001)

**DOI:** 10.1101/2023.08.15.23294144

**Authors:** Yoshinori Katsumata, Yuki Muramoto, Noriyuki Ishida, Ryo Takemura, Kengo Nagashima, Takenori Ikoma, Naoto Kawamatsu, Masaru Araki, Ayumi Goda, Hiroki Okawara, Tomonori Sawada, Yumiko Kawakubo Ichihara, Osamu Hattori, Koki Yamaoka, Yuta Seki, Toshinobu Ryuzaki, Hidehiko Ikura, Daisuke Nakashima, Takeo Nagura, Masaya Nakamura, Kazuki Sato, Yasuyuki Shiraishi

**Affiliations:** Institute for Integrated Sports Medicine, Keio University School of Medicine, Tokyo, Japan; Department of Cardiology, Keio University School of Medicine, Tokyo, Japan; Biostatistics Unit, Clinical and Translational Research Center, Keio University Hospital, Tokyo, Japan; Division of Cardiology, Internal Medicine III, Hamamatsu University School of Medicine, Shizuoka, Japan; Department of Cardiology, Faculty of Medicine, University of Tsukuba, Tsukuba, Japan; Second Department of Internal Medicine, University of Occupational and Environmental Health, Kitakyushu, Japan; Department of Cardiovascular Medicine, Kyorin University Faculty of Medicine, Tokyo, Japan; Department of Orthopaedic Surgery, Keio University School of Medicine, Tokyo, Japan; Department of Clinical Biomechanics, Keio University School of Medicine, Tokyo, Japan

**Keywords:** Heart failure, sweat, anaerobic threshold, ventilatory threshold, lactate threshold, cardiopulmonary exercise testing

## Abstract

**Background:** The ventilatory threshold (VT), which requires an expensive analyzer and expertise in assessment, has not been widely and conveniently used in a clinical setting. This prospective clinical trial (LacS-001) aimed to investigate the safety of a lactate-monitoring sweat sensor, and the correlation between the lactate threshold in sweat (sLT) and the VT, to establish a simple method for determining the anaerobic threshold in patients with heart failure (HF).

**Methods:** We recruited 50 patients with HF and New York Heart Association functional classification I–II, with a median age of 63.5 years (interquartile range: 58.0–72.0 years). Incremental exercise tests were conducted, and changes in sweat lactate levels were monitored simultaneously. sLT was defined as the first steep increase in lactate levels from baseline and was evaluated by three independent investigators. The primary outcome measures were a correlation coefficient of 0.6 or greater between sLT and VT, the similarities as assessed by the Bland–Altman analysis, and the standard deviation of the difference between sLT and VT within 15 W.

**Results:** Among the 50 patients, a correlation coefficient of 0.651 (95% confidence interval, 0.391–0.815) was achieved in 32 cases. The difference between sLT and VT was −4.9±15.0 W, and no comparative error was noted in the Bland–Altman plot. No device-related adverse events were reported among the registered patients.

**Conclusions:** Our sweat lactate sensor provides a safe and accurate means of detecting VT in patients with HF in a clinical setting, thereby offering valuable additional information for treatment.

**Clinical Perspective:** *What is new?:*

- Successful real-time and continuous monitoring of sweat lactate levels with this experimental device during progressive exercise testing in patients with NYHA class I and II HF.
- sLT determined by sweat lactate monitoring correlated strongly with VT, suggesting that VT can be detected with sufficient clinical accuracy.
- Patients with intermediate sweat rates are good candidates for determining sLT.
- Given the difficulties presented by the current methods for determining VT, the monitoring of lactate values in sweat by our sensors could be helpful for improving VT detection.

*What are the clinical implications?:*

- Our sweat lactate sensor provides a safe and accurate means of detecting ventilatory threshold (VT) in patients with heart failure in a clinical setting.
- A correlation coefficient of 0.651 (95% confidence interval, 0.391–0.815) was achieved with −4.9±15.0 W as the difference between the lactate threshold in sweat and VT and no comparative error.
- No device-related adverse events were reported among the registered patients.

## Introduction

Exercise of appropriate quantity and intensity is of paramount importance for good health and the prevention of cardiovascular diseases in all age groups [1,2]. Expert statements and the current clinical practice guidelines recommend aerobic exercise for patients with cardiovascular disease, especially for heart failure (HF) [3]. Anaerobic threshold, also known as ventilatory or lactate threshold, is a key component of exercise intensity used to optimize exercise training programs. An exercise test combined with respiratory gas analysis is necessary to determine ventilatory threshold (VT). However, the exercise test is not fully practical in clinical settings because of the expensive analyzers involved and the required expertise [4].

Recently, interest in wearable sensing technology for a more precise assessment of the physiological responses of the body has notably increased. Human sweat when utilized as a source of physiological information may allow for non-invasive assessments during exercise [5]. To date, sweat lactate sensors are considered impractical alternatives to measuring blood lactate levels because the dynamics of sweat lactate is different from that of blood lactate [6-8]. Therefore, sweat lactate sensors have not been applied in medicine. Considering this, our research group focused on the inflection point (lactate threshold) at which the value increases rapidly during incremental exercise, not the absolute value, and has been verifying the accuracy of this method. We developed a small, easy-to-use device for non-invasive, simple, and real-time visualization of sweat lactate dynamics during exercise [9-14]. This sweat lactate sensor shows increases in sweat lactate levels proportional with fitness intensity. The lactate threshold in sweat (sLT), which is defined as the first steep increase in lactate levels from baseline in sweat, is consistent with VT in healthy individuals and patients with cardiovascular diseases [10]. However, the safety, simplicity of operation, and accuracy of the sensor as a medical device employed in clinical practice need further investigation.

Therefore, the current prospective clinical trial (LacS-001) was designed to investigate the safety of the sweat lactate sensor and to evaluate sLT determination accuracy in patients with HF.

## Methods

### Study design and oversight

The prospective, open-label Lac-001 study (jRCT2032220057) was conducted at Keio University, Japan (see Supplementary material online, *eTable S1*). Patients were recruited between May 2022 and September 2022. The study protocol was approved by the Institutional Review Board of the Keio University School of Medicine (DA21-006). The trial was conducted in accordance with Good Clinical Practice guidelines, the Declaration of Helsinki, and all applicable laws and guidelines in Japan. Patients provided written informed consent before enrolment in the study.

### Study participants

Eligible patients were aged 20–90 years, had a diagnosis of HF, were New York Heart Association (NYHA) class I or II, and planned to exercise. Patients with severe skin disease, tearing at the site where the sensor was applied, or an extremely low perspiration rate (as confirmed by medical interviews) were excluded.

### Endpoints

The primary endpoint was a correlation coefficient of 0.6 or greater between sLT and VT. The similarities among different methods were assessed by the Bland–Altman analysis and the mean and standard deviation (SD) of the difference between the exercise load at VT and sLT. For use in a clinical setting, a difference in the SD of 15 W or less between the exercise load at VT and sLT was considered acceptable for determining the exercise therapy intensity. Using a simulation study, we hypothesized that a correlation coefficient of 0.6 or greater between sLT and VT was required to achieve an SD value within 15 W. The secondary endpoints were 1) the correlation between sLT and VT in clinical practice (VTcp) and 2) interclass correlation coefficient (ICC [2,1]) to investigate inter-evaluator reliability in determining sLT and VTcp. Adverse events were considered any unfavorable medical events occurring within 24 h after the exercise stress test.

### Exercise testing protocol

Exercise tests with respiratory gas analysis were performed by an ergometer with RAMP protocol (10 W or 15 W); simultaneously, a wearable lactate sensor monitored changes in sweat lactate levels. After a 2–3-min calibration period using saline, the sensor chip connected to the monitoring device was attached to the forehead. A perspiration meter (SKN-2000M; SKINOS Co., Ltd., Nagano, Japan) measured the local sweat rate at a sampling rate of 1 Hz in the same area as that of the sweat lactate sensor. Following sensor attachment, participants were tested in an upright position with an electronically braked ergometer (Strength Ergo 8^®^, Mitsubishi Electric Engineering Company, Japan). After a 2-min rest period for stabilization of heart rate and respiratory status, the participants warmed up for 2 min at 0 W and then exercised at progressively increasing intensities until they could no longer maintain the pedaling speed (voluntary fatigue). At 1-min intervals, the intensity increased in increments of 10 or 15 W. The incremental exercise testing time was approximately 10 min based on the exercise capacity of each participant. The pedaling frequency was set to 60 rev/min. Once the exercise tests were terminated, the participants were instructed to stop pedaling and remain on the ergometer for 3 min. The patient’s heart rate was continuously recorded with a 12-lead electrocardiogram. Blood pressure was monitored every minute with an indirect automatic manometer.

### Sweat lactate sensor

Detailed information on the sweat lactate sensor has been previously provided [10-11]. In brief, the wearable device quantifies the lactate concentration through the reaction of lactate dehydrogenase with lactate contained in sweat, which results in a redox reaction that generates an electric current. The current can be acquired as continuous data from 0.1 to 80 µA in 0.1-μA increments. The sweat lactate sensor responded linearly to lactate concentrations, especially in the range of 2–4 mmol/L, which was at the LT [10-11]. The data were continuously recorded on a mobile device via Bluetooth connection at a sampling frequency of 1 Hz. All recorded data were transformed into moving averages at 13-s intervals and individually zero-corrected with baseline levels (Figure 1).

**Figure 1.**
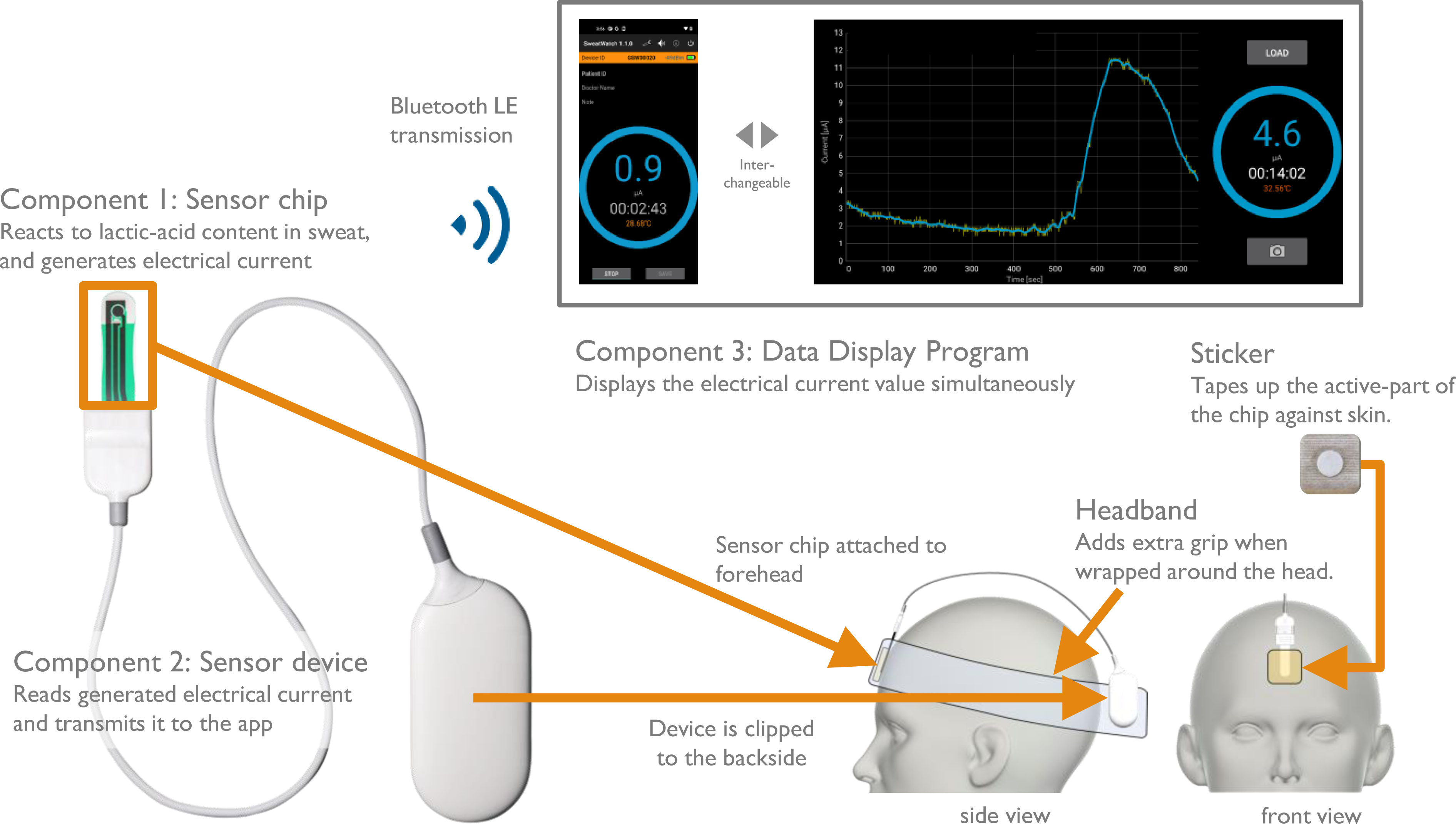
Sweat lactate sensor. The sensor chip connected to the device was attached to the forehead and cleaned with a cloth. The data were recorded continuously at a sampling frequency of 1 Hz for mobile applications using a Bluetooth connection. sLT was defined as the first significant increase in sweat lactate above the baseline, based on graphical plots. sLT, lactate threshold in sweat

### Respiratory gas analysis and VT

The expired gas flows were obtained via a breath-by-breath automated system (Aeromonitor^®^, MINATO MedicalScience CO., LTD., Osaka, Japan). Respiratory gas exchange, such as ventilation output (VE), oxygen uptake (VO_2_), and carbon dioxide generation (VCO_2_), was continuously monitored and averaged at 10 s. A three-way calibration process of a flow volume sensor, a gas analyzer, and a delay-time calibration was performed. Peak VO_2_ was measured as the average oxygen consumption during the last 30 s of exercise. The ventilation/carbon dioxide (ventilator efficiency) slope (VE-VCO_2_ slope) was obtained via linear regression analysis of data from the start of exercise to the respiratory compensation point. Heart rate at rest was defined as the average heart rate 2 min prior to exercise while in the seated position.

VT was assessed using the ventilatory equivalent, excess carbon dioxide, and modified V-slope methods [4,15-17]. In some cases, confirming VT was difficult due to fluctuations in minute ventilation and inconsistencies in several factors, such as VE/VO_2_, terminal exhaled O_2_ concentration, and VCO_2_/VO_2_ slope [18]. Therefore, we investigated the accuracy of sLT using only definite VT (see Supplementary material online, *eFigure S1*). First, VT was independently evaluated by three experienced cardiac rehabilitation experts (physicians) using each of the three methods. If the VTs obtained using three methods were within 15 W, the expert-assessed VT was calculated as the average of the three VTs. If VT was >15 W in one or more of three investigators, it was considered indefinite and excluded from the primary endpoint analysis. Second, if the expert-assessed VTs by two independent investigators were within 5%, VT was determined as the average of the two. If the VTs determined by the independent investigators were not within 5% of one another, the third VT was used for comparison. If the adjudicated VT was within 5% of that reported by either initial investigator, the two VTs were averaged. If the adjudicated VT was within 5% of both values determined by the initial investigators, then the three VTs were averaged. Otherwise, VT was indefinite and excluded from the primary endpoint analysis. Furthermore, each investigator determined VTcp as if testing in a clinical setting while eliminating the false breakpoints in the points assessed by the three methods.

### Lactate threshold in sweat

sLT was defined as the first steep increase on a graphical plot in lactate levels from baseline (Figure 2) and was then evaluated by three investigators who were independent of the VT investigators. Of the three, two did not have cardiac rehabilitation specialist experience; in addition, sLT investigators were not involved in the pilot research related to the sweat lactate sensor [10]. If the sLTs from two investigators were within 5%, they were averaged. If the sLTs of the independent investigators were not within 5% of one another, the third sLT was used for comparison. The two sLTs were averaged when the adjudicated sLT value was within 5% of the values from the initial investigators. If the adjudicated sLT was within 5% of those obtained by both initial investigators, the three sLTs were averaged. Otherwise, sLT was indefinite and excluded from the analysis of the primary endpoint.

**Figure 2.**
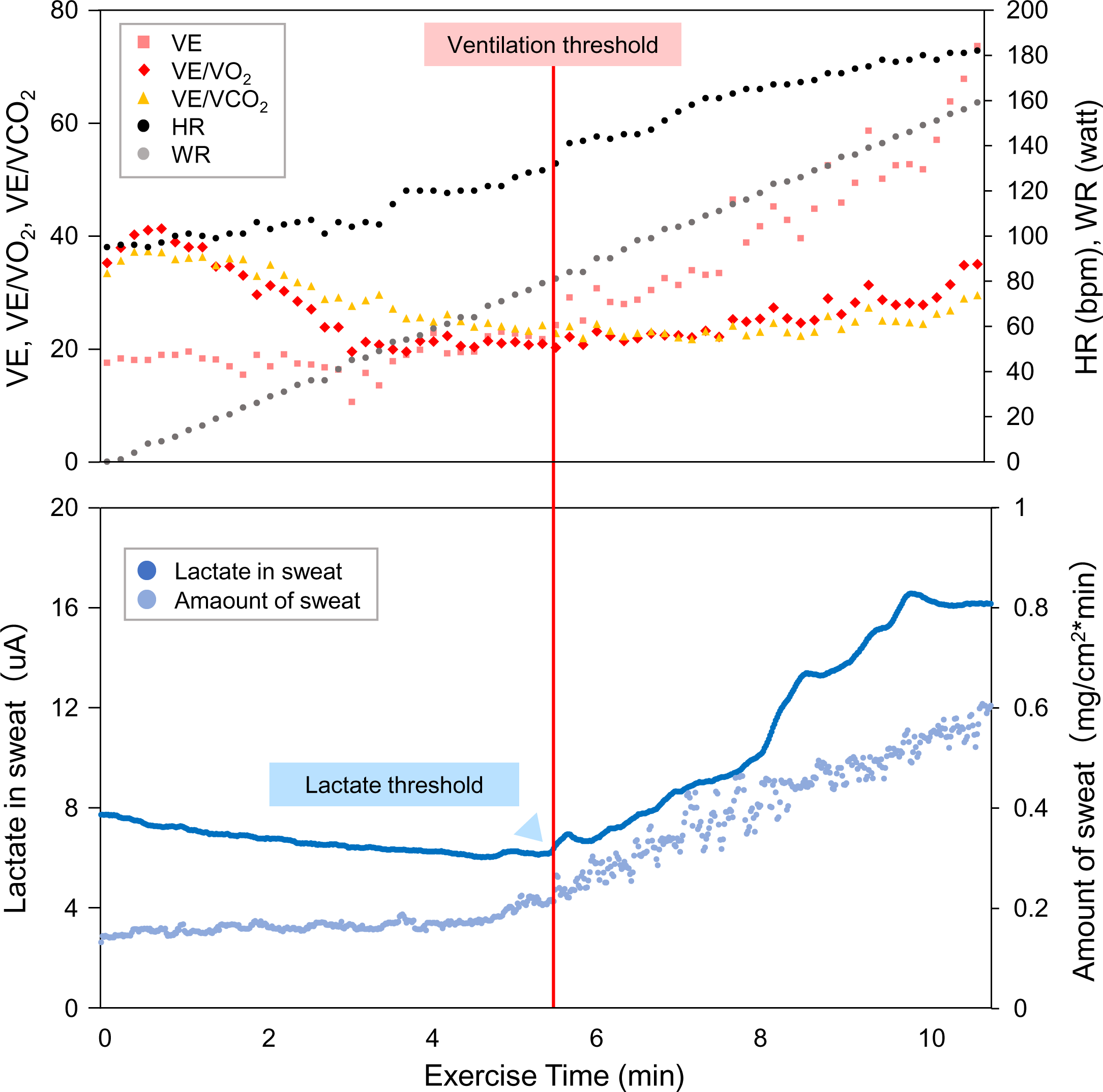
Imaging of sweat lactate levels during incremental exercise. Representative graphs (dots) of lactate in sweat (dark blue) and the amount of sweat produced during exercise using a RAMP (15 W/min) protocol ergometer are shown in the lower panel. Respiratory gas data are shown in the upper panels. HR, heart rate; VE, ventilatory equivalent; VE/VCO_2_, ventilation-carbon dioxide production; VE/VO_2_, ventilation-oxygen uptake; WR, work rate.

### Sample size

A minimum required sample size of 24 patients was needed to achieve statistical significance, assuming an expected correlation coefficient of 0.6 or greater between sLT and VT, with a two-tailed alpha level of 5% and at least 90% power. According to the results of a previous study [10], sLT could not be evaluated in 17% of patients with NYHA class I or II. The VT evaluation had an estimated 30% failure rate, based on clinical experience. In addition to excluding the indefinite thresholds (VT and sLT) from the primary endpoint analysis, the dropout rate was assumed to be 10%. Therefore, a final sample size of 50 was decided.

### Statistical analyses

The results are presented as medians with interquartile ranges (IQR) for continuous variables and as percentages for categorical variables. For the primary endpoint, the relationship between the work rate at sLT and VT was assessed by Pearson’s correlation coefficients. The *P*-value for *H_0_*: *ρ* = 0 and 95% confidence intervals (CI) were calculated. Additionally, the similarities between the different methods were assessed by Bland–Altman analysis [18]. This comparison provides a graphical representation of the differences between the methods and their average values. The mean and SD values of the difference between exercise loads at VT and sLT and 95% CIs were also estimated. For the secondary endpoints, (1) Pearson’s correlation coefficient between sLT and VTcp and (2) ICC [2, 1] were used to investigate the inter-evaluator reliability in determining sLT and VTcp. To evaluate the relationship between sLT determination based on sweat lactate levels and sweat rate, patients were divided into three groups according to the sweat rate tertiles during peak exercise. All analyses were prespecified in a detailed statistical analysis plan that was planned before the first patient enrolment and finalized before the database was locked. All statistical analyses were performed independently and validated by two biostatisticians (NI and RT). All *P*-values were two-tailed; a *P*-value of <0.05 was considered statistically significant. All statistical analyses were performed using the SAS software (version 9.4; SAS Institute, Cary, NC, USA).

## Results

### Study participants

The primary endpoint was met in 32/50 (64.0 %) patients, secondary endpoints in 37/50 (74.0 %), and safety endpoint in 50/50 (100 %) patients (Figure 3). No patient met the exclusion criteria. There were no discontinuations, dropouts, or Good Clinical Practice violations, and 50/50 (100 %) patients completed the study. Of the 50 patients, one was excluded from the primary and secondary analyses due to protocol deviation as the patient underwent an exercise test with a 20-W progressive ergometer. In addition, LT in 12 patients was difficult to determine, as detailed in Supplementary material online, *eTable S2*, and Figure 3. The current was not found to have increased due to the absence of sweat lactate production (see Supplementary material online, *eFigure S2*) or discrepant analysis results between the three independent investigators due to the presence of several inflection points (see Supplementary material online, *eFigure S3*). Therefore, the remaining 37 patients were analyzed for secondary endpoints. Furthermore, VT was difficult to determine in six patients for the reasons stated in Supplementary material online, *eTable S2*, and Figure 3 because of inconsistencies among several factors (such as VE/VO_2_, terminal exhaled O_2_ concentration, and VCO_2_/VO_2_ slope) or discrepant analysis results between the three independent investigators due to the presence of several inflection points (including one patient whose VT and sLT were difficult to assess). In total, the data of 32 patients were analyzed for primary endpoints.

**Figure 3.**
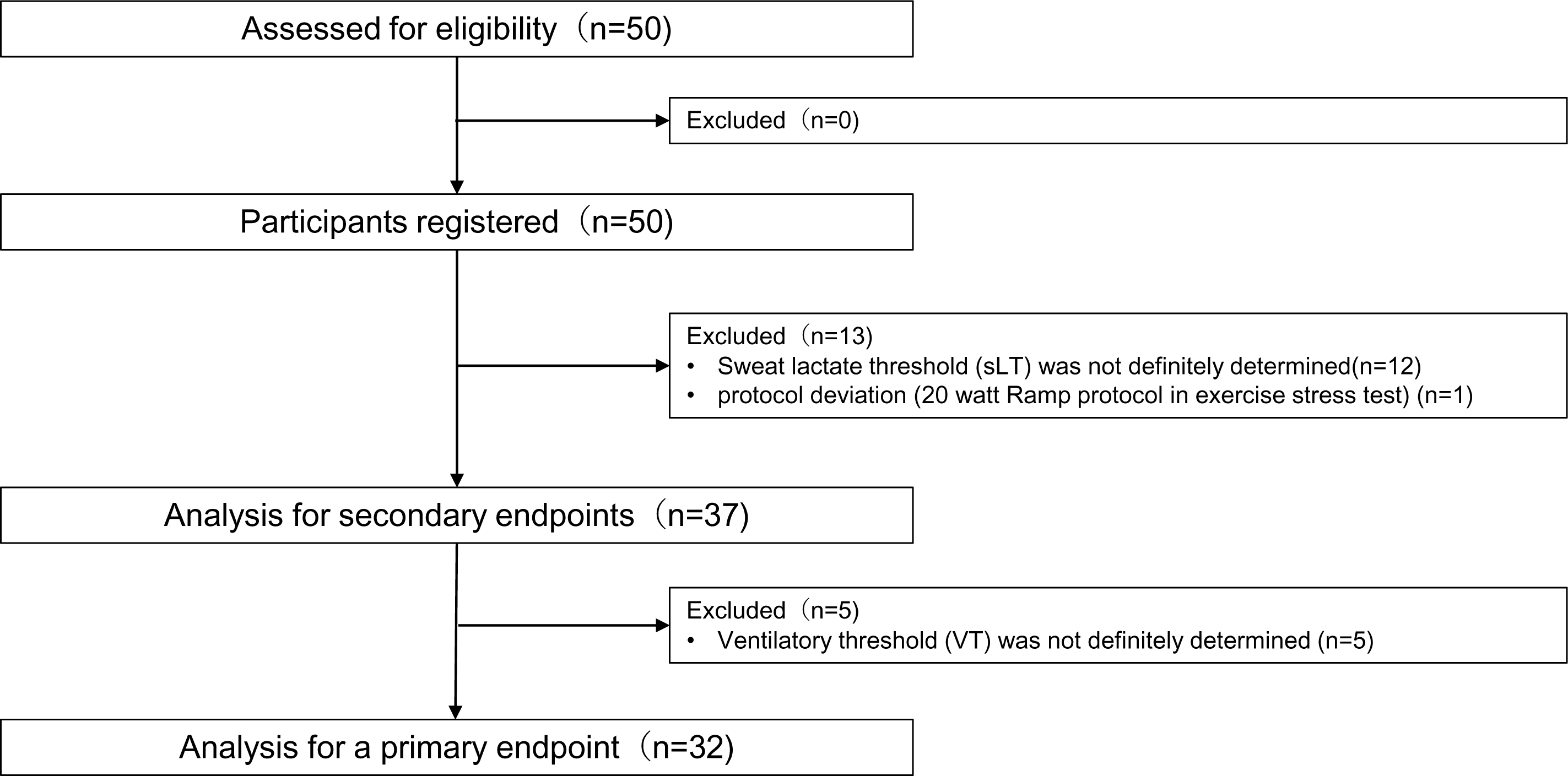
Analysis flow diagram. Of the 50 patients, one patient was excluded from the primary and secondary analyses for protocol deviation because the patient underwent an exercise test with a 20 W progressive ergometer. In addition, LT was difficult to determine in 12 patients. Therefore, the remaining 37 patients were analyzed for secondary endpoints. Furthermore, VT was difficult to determine in six patients. In total, 32 patients were analyzed for primary endpoints. LT, lactate threshold; VT, ventilatory threshold.

The baseline characteristics are summarized in Table 1 and Supplementary material online, *eTable S3*. The patients were predominantly male (88%) with a median age of 63.5 years (IQR, 58.0–72.0) and left ventricular ejection fraction of 59.6% (IQR, 51.6–64.0). Thirty-four patients (84%) were on beta-blockers. Two, six, and one patients were treated with a pacemaker, an implantable cardioverter defibrillator, and cardiac resynchronization therapy, respectively. The proportions of patients with NYHA functional classes I and II were 60% and 40%, respectively. Forty-four (88%), one (2%), and five (10%) patients had sinus rhythm, atrial fibrillation, and pacemaker rhythm, respectively. The cardiopulmonary exercise test results are shown in Supplementary material online, *eTable S4, 5, and 6*. All patients completed the exercise tests without complications; testing was discontinued when patients could no longer maintain the indicated pedal rate. The peak respiratory quotient was 1.18. Among the cardiopulmonary exercise test variables, peak VO_2_, peak heart rate, and VE/VCO_2_ slope were 20.1 mL/kg/min (16.7–24.0), 127.5/min (115.0–144.0), and 29.8 (26.0–33.6), respectively.

**Table 1.**
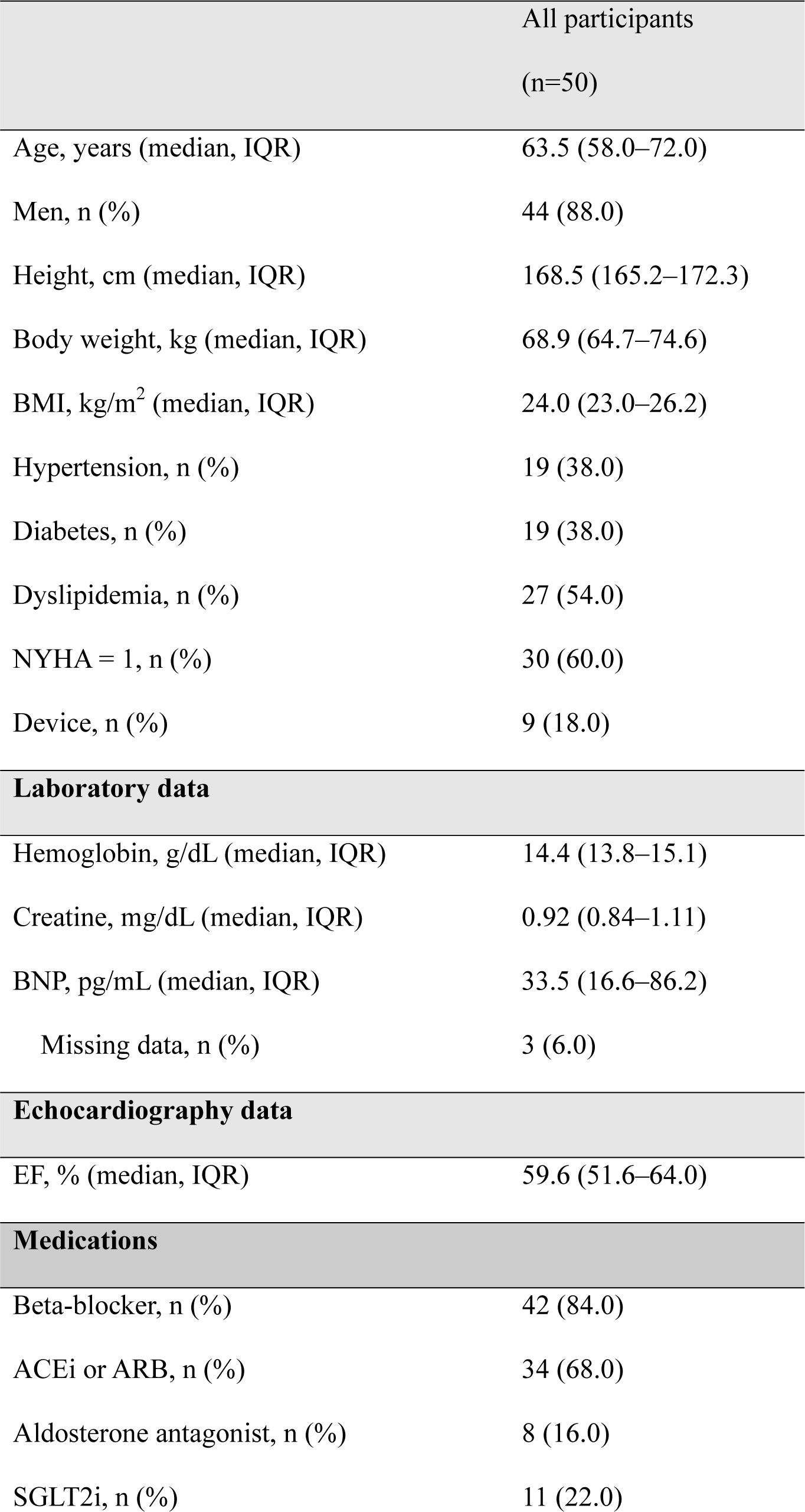
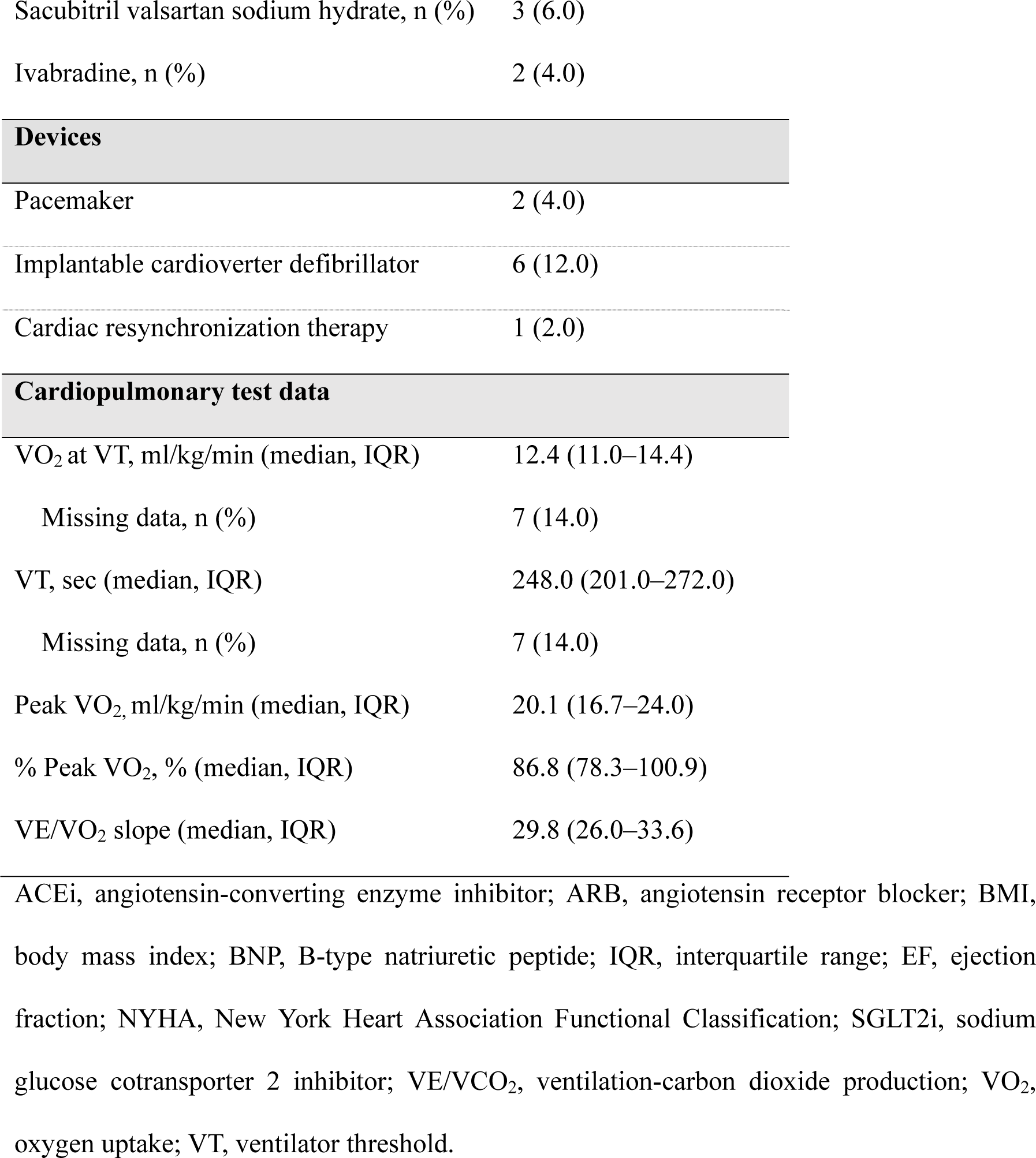
Baseline characteristics.

### Monitoring the lactate levels in sweat during exercise

Figure 2 shows the sweat lactate values during progressive exercise tests. Real-time changes in sweat lactate levels were immediately reflected on the wearable device. At the commencement of cycling activity, the current response from the lactate sensor was negligible because of the lack of sweat. After the onset of sweating, lactate released from the epidermis was selectively detected by the sensor. During exercise, sweat lactate values continued to increase until volitional exhaustion. After the exercise period, a relatively slow decrease in sweat lactate values was observed compared with the decrease in the heart rate. The sweat rate was 0.40 mg/min/cm^2^ (IQR, 0.20–0.60 mg/min/cm^2^) during peak exercise. When the patients were divided into three groups by tertiles of sweat rate in peak exercise, the sweat rate of each group (low, intermediate, and high) was 0.16 mg/min/cm^2^ (IQR, 0.07– 0.20 mg/min/cm^2^), 0.43 mg/min/cm^2^ (IQR, 0.31–0.48 mg/min/cm^2^), and 0.66 mg/min/cm^2^ (IQR, 0.61–1.09 mg/min/cm^2^), respectively (Figure 4A). The frequencies of cases in which sLT could definitely be determined at low, intermediate, and high sweat rates were 53%, 100%, and 75%, respectively (Figure 4B).

**Figure 4.**
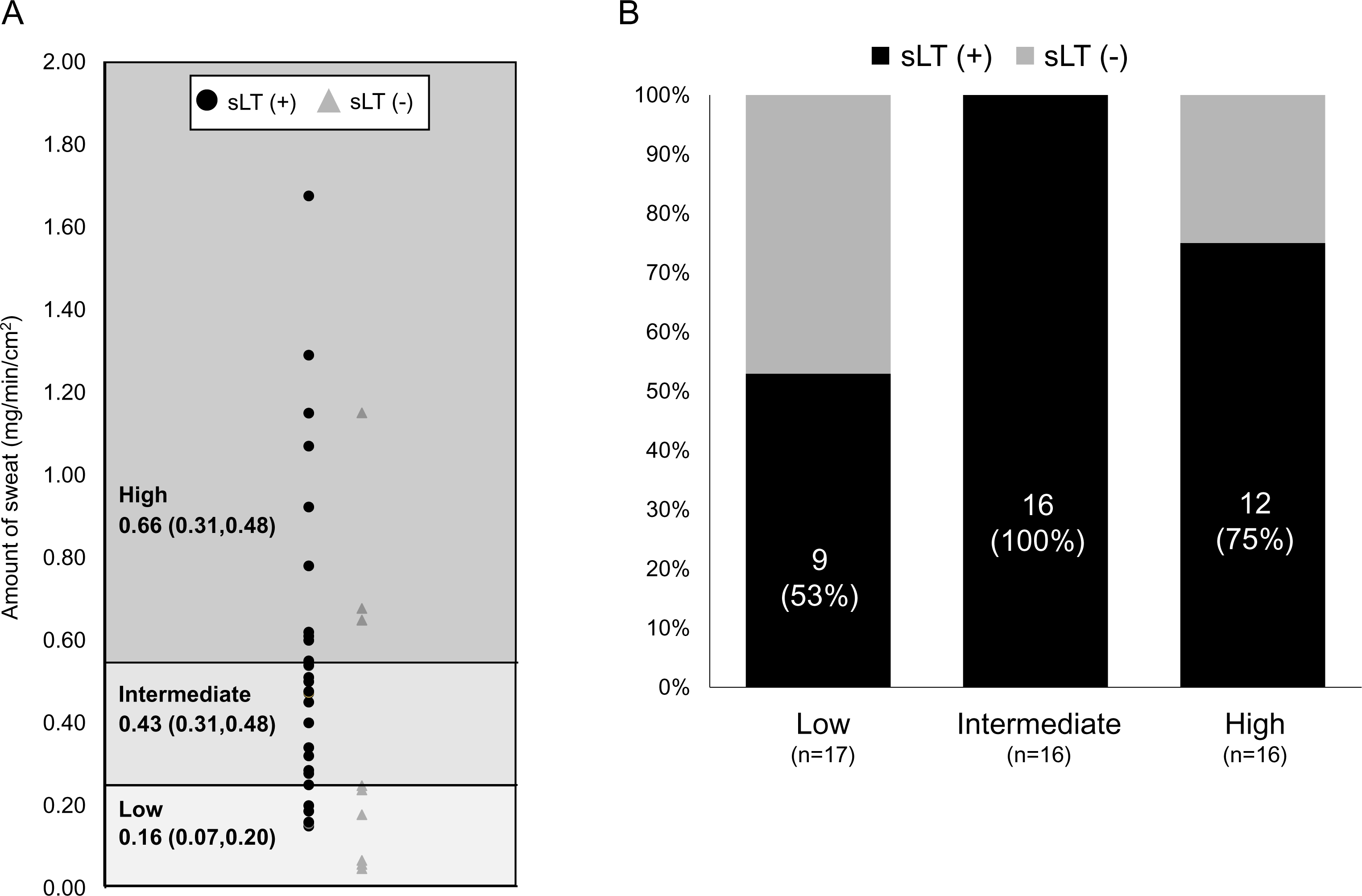
Frequency of patients with definitely determined sweat lactate thresholds based on the amount of sweat. The patients were divided into three groups according to the tertiles of the amount of sweat produced during peak exercise. A) Black circles indicate patients with a definite sweat lactate threshold (sLT+), and navy triangles indicate patients with no definite sweat lactate threshold (sLT-). Sweat rate during peak exercise of each group was 0.16 mg/min (IQR, 0.07–0.20), 0.43 mg/min (IQR, 0.31–0.48), and 0.66 mg/min (IQR, 0.61–1.09). B) Frequency of cases in which the sweat lactate threshold was determined at low, intermediate, and high sweat rates. sLT, lactate threshold in sweat.

### Primary endpoint—the relationship between sLT and VT

A strong correlation was observed between sLT and VT (r=0.651, 95% CI: 0.391–0.815, *P*<0.001; Figure 5A and Table 2). The Bland–Altman plot showed a nearly symmetrical distribution centered around 0, with no proportional error (Figure 5B; mean difference between each threshold: −4.9±15.0 W).

**Figure 5.**
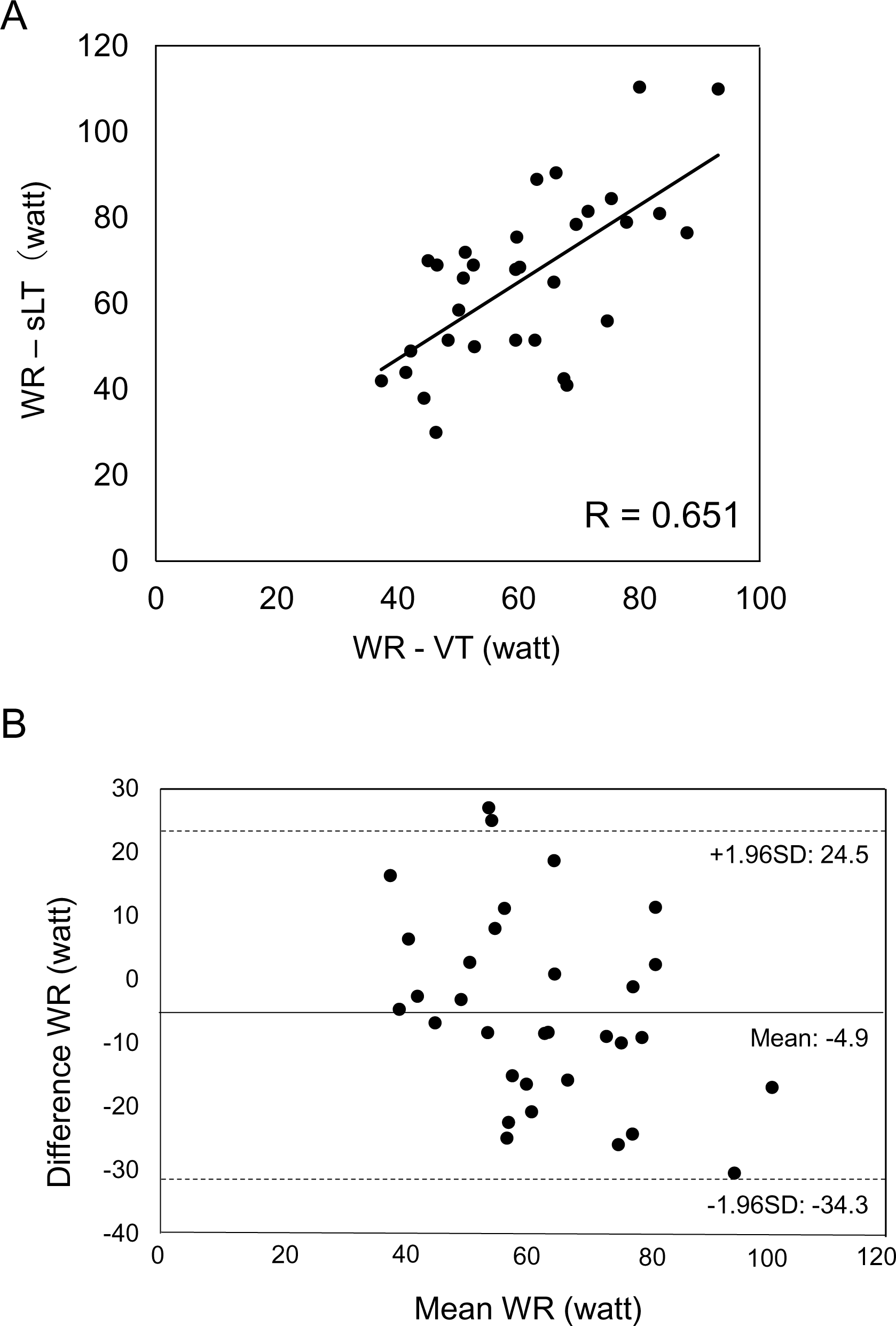
Validity testing of the work rate at the ventilatory and sweat lactate thresholds. A) The relationship between the work rate (WR) at the lactate threshold in sweat (WR-sLT) and the WR-ventricular threshold (WR-VT). (B) The Bland–Altman plots, which indicate the respective differences between WR-sLT and WR-VT (y-axis) for each individual against the mean of WR-sLT and WR-VT1 (x-axis). R indicates the correlation coefficient; SD, standard deviation; sLT, lactate threshold in sweat; WR, work rate; VT, ventilatory threshold

**Table 2.**
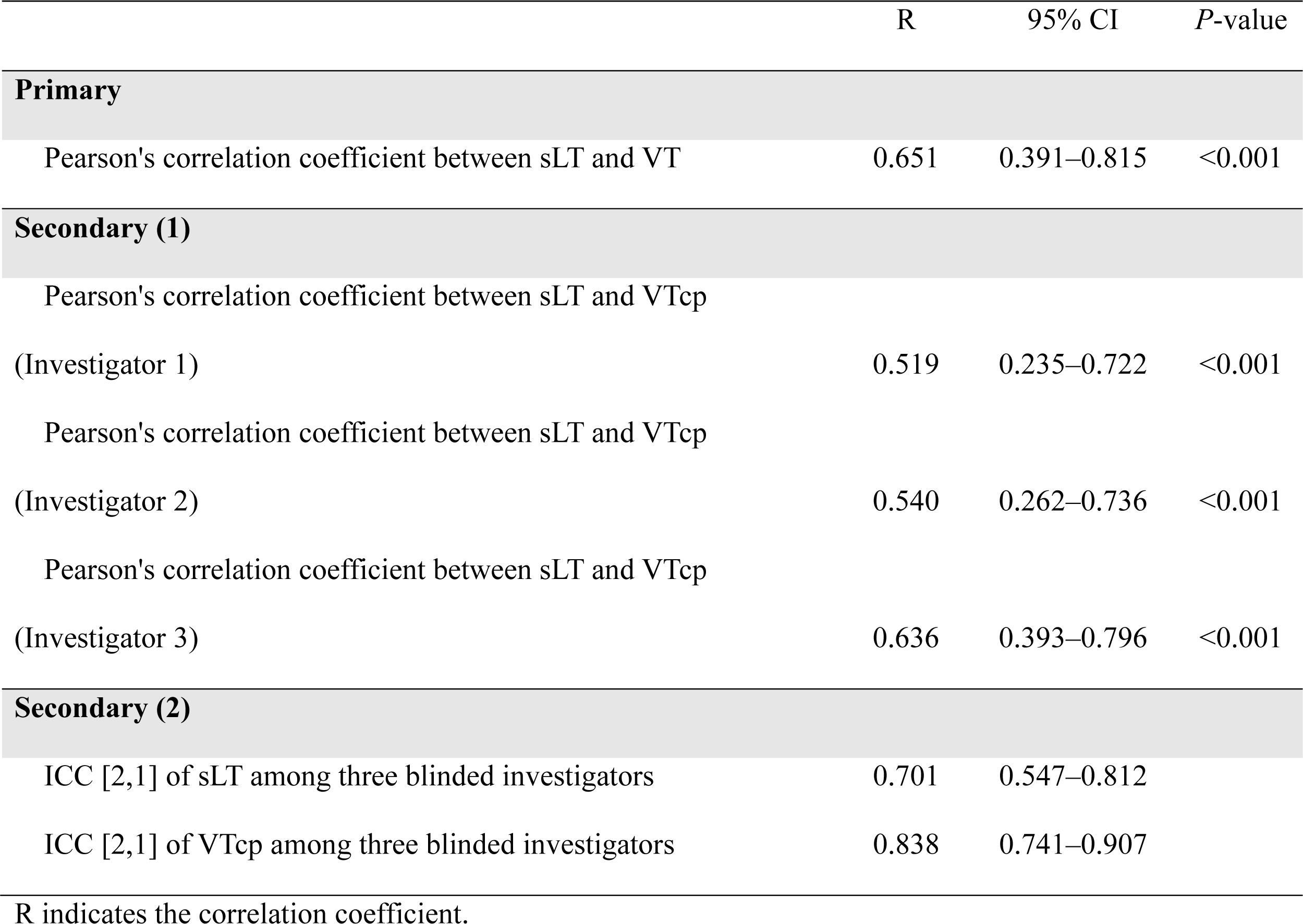
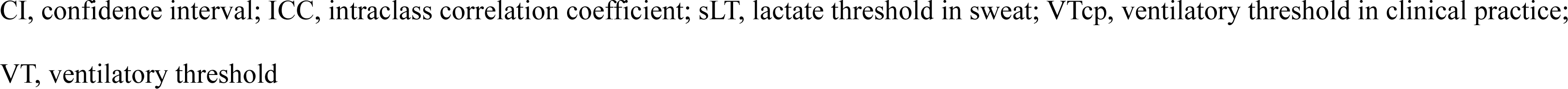
Primary and secondary endpoints.

|  | R | 95% CI | P-value |
| --- | --- | --- | --- |
| **Primary** |  |  |  |
| Pearson's correlation coefficient between sLT and VT | 0.651 | 0.391–0.815 | <0.001 |
| **Secondary (1)** |  |  |  |
| Pearson's correlation coefficient between sLT and VTcp |  |  |  |
| (Investigator 1) | 0.519 | 0.235–0.722 | <0.001 |
| Pearson's correlation coefficient between sLT and VTcp |  |  |  |
| (Investigator 2) | 0.540 | 0.262–0.736 | <0.001 |
| Pearson's correlation coefficient between sLT and VTcp |  |  |  |
| (Investigator 3) | 0.636 | 0.393–0.796 | <0.001 |
| **Secondary (2)** |  |  |  |
| ICC [2,1] of sLT among three blinded investigators | 0.701 | 0.547–0.812 |  |
| ICC [2,1] of VTcp among three blinded investigators | 0.838 | 0.741–0.907 |  |

R indicates the correlation coefficient.
CI, confidence interval; ICC, intraclass correlation coefficient; sLT, lactate threshold in sweat; VTcp, ventilatory threshold in clinical practice; VT, ventilatory threshold

### Secondary endpoints

Regarding the secondary endpoint (1), strong correlations were also observed between the sLT and VTcp determined by the three blinded investigators (investigator 1: r=0.519, 95% CI, 0.235–0.722; investigator 2: r=0.540, 95% CI, 0.262–0.736; investigator 3: r=0.636, 95% CI, 0.393–0.796; Table 2). Regarding the secondary endpoint (2), both sLT and VTcp showed very good interrater reliability among the three blinded investigators (measured value [95% CI]: sLT, 0.701 [0.547–0.812]; VTcp, 0.838 [0.741–0.907]; Table 2).

### Safety of the exercise test using the sweat lactate sensor

During the exercise stress test, the adverse events of ventricular arrhythmia or tachycardia occurred in two patients. These events were caused by exercise stress and were unrelated to the lactate sensor. No serious adverse events were associated with the sensor.

## Discussion

In patients with NYHA class I or II HF, it was possible to measure lactate levels in sweat during a progressive exercise stress test using a lactate sensor in a continuous and real-time manner. A strong correlation was observed between sLT and VT, suggesting that VT can be detected by real-time monitoring of sweat lactate levels with sufficient accuracy for clinical use. The recommended exercise intensity for patients with HF is 40–60% of the peak oxygen intake [19,20]. In addition, exercise was performed using heart rate in combination with the Borg scale based on VT or sLT. Therefore, we hypothesized that sLT determination using the device would be clinically acceptable if the SD value of the difference between sLT and VT was approximately within 15 W, and our results were within that range. Two adverse events occurred in this study: both were arrhythmias (supraventricular tachycardia and ventricular tachycardia) during exercise testing. These events were caused by exercise stress and were unrelated to the lactate sensor. Furthermore, no adverse events such as redness, swelling of the skin contact areas, or serious adverse events were observed.

The biosensor must be flexible enough to adhere to the placement site without disturbing the user. Recent advances in manufacturing technology have allowed for the design of biosensors capable of providing a natural fit to the smooth curved geometry of human skin while maintaining suitable contact for the physiological measurement of chemicals and electrolytes [21]. Polyethylene terephthalate substrates were used to create a highly flexible sensor that is easily aligned with curved surfaces. The upper arm and forehead are appropriate sites for monitoring lactate levels in sweat because of their smooth skin surfaces for sensor placement and non-interference during pedaling tasks [9-14]. Particularly in patients with HF who exhibit insufficient sweating due to old age or drugs such as diuretics, the forehead has been used by our team because of the high sweat rate in this area during exercise [9,22].

During physical activity, such as exercise, complex physiological changes have been observed in the human body [23]. Current wearable devices only obtain physical or electrophysiological information, such as heart rate, exercise, intermittent blood pressure, arterial oxygen saturation, and electrocardiogram; however, advances in sensing modalities are expected to help obtain a detailed picture of the complex physiological changes that occur during exercise [24-26]. The continuous measurement of lactate levels during exercise can provide useful insights into physiological changes that are otherwise not possible to obtain based on physical or electrophysiological information [27]. In particular, the concentration of blood lactate has been used to monitor individual performance and exercise intensity [27-29]. Currently, blood lactate levels are measured in clinical laboratories or by using point-of-care devices; however, this approach does not allow for continuous real-time measurements. In contrast, our device, which utilizes sweat on the skin, successfully measured real-time sweat lactate concentrations continuously and noninvasively during progressive exercise testing in patients with NYHA class I or II HF, which allows for the visualization of complex physiological changes during exercise.

A pilot study suggested that many patients with NYHA class III or higher and a lower maximal oxygen uptake had sweat lactate levels below the sensitivity of the measurement method [10]. To enroll patients with sufficient sweat production for a lactate sensor, those with NYHA class I or II and no history of low sweating rates were eligible. However, in 13 patients, sLT was definitely not determined. It is not surprising that the sensor using sweat cannot be utilized for all patients due to the diversity of sweat dynamics during exercise, including sweat volume [30]. Since it is clinically routine to perform exercise stress tests in patients with HF in practice while monitoring heart rate and Borg scale scores, patients for whom our sensor cannot determine sLT will be presented with an exercise prescription using these parameters. Detailed analysis showed that sLT could be determined in all patients with intermediate levels of sweating (median, 0.43 mg/min/cm^2^ [IQR, 0.31–0.48 mg/min/cm^2^] in sweating during peak exercise). Patients with insufficient sweating during exercise did not respond to the lactate sensor; thus, sLT could not be determined. Additionally, some of the patients with a high sweating rate had no inflection point or few candidate points, resulting in an inability to steeply refine the sweat lactate level to a single point. These findings indicate that patients with intermediate sweat rates are good candidates for determining sLT using our sensors. Adjustment of the exercise environment (e.g., humidity and temperature) and duration (e.g., a long warm-up and total exercise time) may be required for patients with low or high sweat rates. In addition, hermetic sealing or high local temperatures that promote sweating can overcome issues related to insufficient sweating in normal environments. The use of a respiratory gas analyzer has the possibility of cross-infection because of the closed circuit. The determination of sLT by sweat-based monitoring has the potential to overcome these problems. Considering the low rate of outpatient cardiac rehabilitation (7%) [31], and the small number of facilities with breath gas analyzers, our sensor could significantly contribute to the cardiac rehabilitation of patients with HF with sufficient accuracy.

Our findings should be interpreted with the following limitations in mind. Although this study was designed for NYHA class I and II HF, patients undergoing cardiac rehabilitation are also indicated for NYHA III. Therefore, further studies are needed to examine which patients with HF are indicated for this sensor. Many participants have relatively high cardiac function and exercise tolerance; thus, there is an urgent need to design a study protocol or develop sensors for use in patients with severe HF. In addition, most patients enrolled in this study were men, which does not reflect the general HF population accurately. Thus, the efficacy and safety of this sensor in women should also be verified. The form of exercise loading used in this study was limited to the use of an ergometer; thus, it is unclear whether the sensor can be adapted to exercise testing using a treadmill.We succeeded in real-time and continuous monitoring of sweat lactate levels using this experimental device during progressive exercise testing in patients with NYHA class I and II HF. sLT determined by sweat lactate monitoring correlated strongly with VT, suggesting that VT can be detected with sufficient clinical accuracy. Our findings also indicated that patients with intermediate sweat rates are good candidates for determining sLT. Given the difficulties presented by the current methods for determining VT, the monitoring of lactate values in sweat by our sensors could be helpful for improving VT detection.

## Acknowledgments

The authors thank C. Yoshida, K. Takeuchi, W. Seki, and Y. Yonezawa for their technical assistance (Keio University School of Medicine). The authors also thank CMIC CO. Ltd. for their supportive study management. We would like to thank Editage (www.editage.com) for English language editing.

## Data availability

The datasets used and/or analyzed in the current study are available from the corresponding author upon reasonable request.

## Non-standard Abbreviations and Acronyms

CI: confidence interval
HF: heart failure;
ICC: interclass correlation coefficient;
NYHA: New York Heart Association;
SD: standard deviation;
sLT: lactate threshold in sweat;
VCO2: carbon dioxide generation;
VE: ventilation output;
VO_2_: oxygen uptake;
VT: ventilator threshold

## References

[1] Schnohr P, O’Keefe JH, Marott JL, Lange P, Jensen GB. Dose of jogging and long-term mortality: the Copenhagen City Heart Study. J Am Coll Cardiol. 2015;65(5):411–419. doi:10.1016/j.jacc.2014.11.023.

[2] Wen CP, Wai JP, Tsai MK, et al. Minimum amount of physical activity for reduced mortality and extended life expectancy: a prospective cohort study. Lancet. 2011;378(9798):1244–1253. doi:10.1016/S0140-6736(11)60749-6.

[3] Belardinelli R, Georgiou D, Cianci G, Purcaro A. 10-year exercise training in chronic heart failure: a randomized controlled trial. J Am Coll Cardiol. 2012;60(16):1521–1528. doi:10.1016/j.jacc.2012.06.036.

[4] Binder RK, Wonisch M, Corra U, et al. Methodological approach to the first and second lactate threshold in incremental cardiopulmonary exercise testing. Eur J Cardiovasc Prev Rehabil. 2008;15(6):726–734. doi:10.1097/HJR.0b013e328304fed4.

[5] Baker LB. Sweating Rate and Sweat Sodium Concentration in Athletes: A Review of Methodology and Intra/Interindividual Variability. Sports Med. 2017;47(Suppl 1):111–128. doi:10.1007/s40279-017-0691-5.

[6] Baker LB, Wolfe AS. Physiological mechanisms determining eccrine sweat composition. Eur J Appl Physiol. 2020;120(4):719–752. doi:10.1007/s00421-020-04323-7.

[7] Green JM, Bishop PA, Muir IH, McLester JR Jr, Heath HE. Effects of high and low blood lactate concentrations on sweat lactate response. Int J Sports Med. 2000;21(8):556–560. doi:10.1055/s-2000-8483.

[8] Lamont LS. Sweat lactate secretion during exercise in relation to women’s aerobic capacity. J Appl Physiol (1985). 1987;62(1):194–198. doi:10.1152/jappl.1987.62.1.194.

[9] Katsumata Y, Sano M, Okawara H, et al. Laminar flow ventilation system to prevent airborne infection during exercise in the COVID-19 crisis: A single-center observational study. PLoS One. 2021;16(11):e0257549. Published 2021 Nov 10. doi:10.1371/journal.pone.0257549.

[10] Seki Y, Nakashima D, Shiraishi Y, et al. Anovel device for detecting anaerobic threshold using sweat lactate during exercise. Sci Rep. 2021 Mar 2;11(1):4929. doi: 10.1038/s41598-021-84381-9.

[11] Muramoto Y, Nakashima D, Amano T, et al. Estimation of maximal lactate steady state using the sweat lactate sensor.Sci Rep. 2023 Jun 26;13(1):10366. doi: 10.1038/s41598-023-36983-8.

[12] Okawara H, Sawada T, Nakashima D, et al. Kinetic changes in sweat lactate following fatigue during constant workload exercise. Physiol Rep. 2022;10(2):e15169. doi:10.14814/phy2.15169.

[13] Sawada T, Okawara H, Nakashima D, et al. Constant Load Pedaling Exercise Combined with Electrical Muscle Stimulation Leads to an Early Increase in Sweat Lactate Levels. Sensors (Basel). 2022;22(24):9585. Published 2022 Dec 7. doi:10.3390/s22249585.

[14] Maeda Y, Okawara H, Sawada T, et al. Implications of the Onset of Sweating on the Sweat Lactate Threshold. Sensors (Basel). 2023;23(7):3378. Published 2023 Mar 23. doi:10.3390/s23073378.

[15] Gaskill SE, Ruby BC, Walker AJ, Sanchez OA, Serfass RC, Leon AS. Validity and reliability of combining three methods to determine ventilatory threshold. Med Sci Sports Exerc. 2001;33(11):1841–1848. doi:10.1097/00005768-200111000-00007.

[16] Mezzani A, Agostoni P, Cohen-Solal A, et al. Standards for the use of cardiopulmonary exercise testing for the functional evaluation of cardiac patients: a report from the Exercise Physiology Section of the European Association for Cardiovascular Prevention and Rehabilitation. Eur J Cardiovasc Prev Rehabil. 2009;16(3):249–267. doi:10.1097/HJR.0b013e32832914c8.

[17] Mezzani A, Hamm LF, Jones AM, et al. Aerobic exercise intensity assessment and prescription in cardiac rehabilitation: a joint position statement of the European Association for Cardiovascular Prevention and Rehabilitation, the American Association of Cardiovascular and Pulmonary Rehabilitation and the Canadian Association of Cardiac Rehabilitation. Eur J Prev Cardiol. 2013;20(3):442–467. doi:10.1177/2047487312460484.

[18] Bland JM, Altman DG. Statistical methods for assessing agreement between two methods of clinical measurement. Lancet. 1986;1(8476):307–310..

[19] Nishi I, Noguchi T, Furuichi S, et al. Are cardiac events during exercise therapy for heart failure predictable from the baseline variables?. Circ J. 2007;71(7):1035–1039. doi:10.1253/circj.71.1035.

[20] Fernhall B. Long-term aerobic exercise maintains peak VO(2), improves quality of life, and reduces hospitalisations and mortality in patients with heart failure. J Physiother. 2013;59(1):56. doi:10.1016/S1836-9553(13)70149-8.

[21] D.H. Kim, N. Lu, R. Ma, et al, Epidermal electronics, Science. 32 (2014) 838–843.

[22] Havenith G, Fogarty A, Bartlett R, Smith CJ, Ventenat V. Male and female upper body sweat distribution during running measured with technical absorbents. Eur J Appl Physiol. 2008;104(2):245–255. doi:10.1007/s00421-007-0636-z.

[23] Tatterson AJ, Hahn AG, Martin DT, Febbraio MA. Effects of heat stress on physiological responses and exercise performance in elite cyclists. J Sci Med Sport. 2000;3(2):186–193. doi:10.1016/s1440-2440(00)80080-8.

[24] Bandodkar AJ, Wang J. Non-invasive wearable electrochemical sensors: a review. Trends Biotechnol. 2014;32(7):363–371. doi:10.1016/j.tibtech.2014.04.005.

[25] Gao W, Brooks GA, Klonoff DC. Wearable physiological systems and technologies for metabolic monitoring. J Appl Physiol (1985). 2018;124(3):548-556. doi:10.1152/japplphysiol.00407.2017.

[26] Wen L, Xie D, Wu J, et al. Humidity-/Sweat-Sensitive Electronic Skin with Antibacterial, Antioxidation, and Ultraviolet-Proof Functions Constructed by a Cross-Linked Network. ACS Appl Mater Interfaces. 2022;14(50):56074–56086. doi:10.1021/acsami.2c15876.

[27] Urhausen A, Coen B, Weiler B, Kindermann W. Individual anaerobic threshold and maximum lactate steady state. Int J Sports Med. 1993;14(3):134–139. doi:10.1055/s-2007-1021157.

[28] Beneke R. Methodological aspects of maximal lactate steady state-implications for performance testing. Eur J Appl Physiol. 2003;89(1):95–99. doi:10.1007/s00421-002-0783-1.

[29] Faude O, Kindermann W, Meyer T. Lactate threshold concepts: how valid are they?. Sports Med. 2009;39(6):469–490. doi:10.2165/00007256-200939060-00003.

[30] Buono MJ, Lee NV, Miller PW. The relationship between exercise intensity and the sweat lactate excretion rate. J Physiol Sci. 2010;60(2):103–107. doi:10.1007/s12576-009-0073-3.

[31] Kamiya K, Yamamoto T, Tsuchihashi-Makaya M, et al. Nationwide Survey of Multidisciplinary Care and Cardiac Rehabilitation for Patients With Heart Failure in Japan - An Analysis of the AMED-CHF Study. Circ J. 2019;83(7):1546–1552. doi:10.1253/circj.CJ-19-0241.

